# Relationship between intragastric meal distribution, gastric emptying and gastric neuromuscular dysfunction in chronic gastroduodenal disorders

**DOI:** 10.1101/2025.02.04.25321689

**Authors:** Chris Varghese, Armen A Gharibans, Daphne Foong, Gabriel Schamberg, Stefan Calder, Vincent Ho, Reena Anand, Christopher N Andrews, Alan H Maurer, Thomas Abell, Henry P Parkman, Greg O’Grady

**Author notes:** **Corresponding Author**, Prof. Greg O’Grady, Department of Surgery, University of Auckland, New Zealand. **Disclosures:** AG, CA, TA, and GO hold grants and intellectual property in the field of GI electrophysiology. GO, AG, GS, SC, CA, DF, and CV are members of the University of Auckland spin-out companies: The Insides Company (GO), and Alimetry (AG, DF, GS, SC, CV, CA, and GO). AM, TA, HP, and RA have no conflicts of interest to declare.

## Abstract

**Background:** Chronic gastroduodenal symptoms arise from heterogenous gastric motor dysfunctions. This study applied multimodal physiological testing using gastric emptying scintigraphy (GES) with intragastric meal distribution (IMD) and Gastric Alimetry® body surface gastric mapping (BSGM) to define motility and symptom associations.

**Methods:** Patients with chronic gastroduodenal symptoms underwent simultaneous supine GES and BSGM with 30 m baseline, 99mTC-labelled egg meal, and 4 h postprandial recording. IMD (ratio of counts in the proximal half of the stomach to the total gastric counts) was calculated immediately after the meal (IMD0), with <0.568 defining impaired accommodation. BSGM phenotyping followed a consensus approach, based on normative spectral reference intervals.

**Results:** Among 67 patients (84% female, median age 40, median BMI 24), median IMD0 was 0.76 (IQR 0.69-0.86) with 5 (7.5%) meeting impaired accommodation criteria. Delayed gastric emptying (n=18) was associated with higher IMD0 (median 0.9 vs 0.7, p=0.004). On BSGM, 15 patients had abnormal spectrograms (5 [7.5%] high frequency and 10 (14.9%) low rhythm stability and/or amplitude); and in these patients, higher IMD0 (proximal retention) strongly correlated to delayed BSGM meal responses (R=-0.71, p=0.003). Lower IMD, indicating antral distribution, correlated with higher gastric frequencies (R=-0.27, p=0.03). BSGM abnormalities paired with impaired accommodation were associated with worse dyspeptic symptoms.

**Conclusion:** Proximal retention of food as assessed by intragastric meal distribution correlated with delayed emptying, and in the presence of neuromuscular spectral abnormalities (abnormal frequencies or rhythms), delayed motility responses on BSGM. Patients with multiple motor abnormalities experience worse dyspeptic symptoms.

## Introduction

Chronic neurogastroduodenal disorders arise from heterogenous mechanisms, with some patients demonstrating motor dysfunction, and others seemingly lacking motility abnormalities. Whilst a range of motility abnormalities such as impaired accommodation, antral hypomotility, pyloric dysfunction are implicated in chronic gastroduodenal disorders,^1,2^ non-motility related factors such as inflammation, visceral hypersensitivity, hormonal feedback loops, and centrally-mediated mechanisms also contribute.^3^ Robustly defining these features may help clarify diagnostic classifications and guide personalized therapies.

Gastric emptying testing has been the most widely used test of gastric function, with scintigraphy being the gold-standard for evaluating gastric transit. However, gastric emptying status correlates poorly with symptoms and can be labile over time.^4,5^ Additional to summative measures of transit, scintigraphic assessment of gastric emptying can allow evaluation of meal distributions when advanced analysis methods are applied, offering a non-invasive biomarker of fundic accommodation.^6^ Impairments of intragastric meal distributions measured in this way have previously been implicated in functional dyspepsia.^7,8^

Recently, Gastric Alimetry has emerged as a new non-invasive test of gastric myoelectrical activity, enabling robust evaluation of the myoelectrical coordination of gastric motility,^9,10^ and the dynamic postprandial meal response.^11^ In addition, validated time-of-test symptom profiling aids comparison of physiological abnormalities as they pertain to symptom genesis.^10,12^

The aim of this study was therefore to apply multimodal physiological profiling using gastric emptying scintigraphy (GES) with intragastric meal distribution (IMD) together with Gastric Alimetry® body surface gastric mapping (BSGM) to comprehensively define motility and symptom associations in patients with chronic gastroduodenal disorders.

## Methods

This was a prospective observational cohort study conducted in Auckland, New Zealand and Western Sydney, Australia. Ethical approvals were granted at each institution (AHREC123; H13541), and all patients provided informed consent. The study is reported in accordance with the STROBE statement.^13^

### Inclusion criteria

Consecutive patients aged ≥18 years with chronic gastroduodenal symptoms and negative upper gastrointestinal endoscopy undergoing GES were invited to participate. Exclusion criteria included those with known structural gastrointestinal diseases and previous abdominal surgery. Patients with cyclic vomiting syndrome or cannabinoid hyperemesis were also excluded. Specific exclusion criteria for Gastric Alimetry including BMI of >35, active abdominal wounds or abrasions, fragile skin, and allergies to adhesives were also applied.

### Physiological testing

Patients underwent simultaneous Gastric Alimetry (Alimetry, New Zealand) and GES testing using standardized test protocols.^10,14^ Tests were conducted after an overnight fast, and medications affecting GI motility were withheld for 48 h prior to testing. Patients were asked to avoid caffeine, nicotine, opiates, and cannabis on the day of testing. Selective serotonin reuptake inhibitors were withheld on the morning of testing. Glucose levels were controlled in diabetic subjects. The standardized ∼255 kCal low-fat egg meal radiolabeled with ^99m^Tc was used, as previously reported.^14,15^ The egg meal is known to elicit the same meal response as the standard Gastric Alimetry meal, however it is noted that amplitude and the Gastric Alimetry Rhythm Index (GA-RI) can be lower than seen with the larger standard Gastric Alimetry oatmeal and nutrient drink (482 kCal) meal.^16^

#### Gastric emptying scintigraphy

GES images were acquired at 0, 1, 2, and 4 h after meal ingestion whilst patients’ were in supine position. Delayed gastric emptying was present if gastric retention was >10% at 4 h.^14^ IMD is the ratio of counts in the proximal half of the stomach to the total gastric counts which was calculated immediately after the meal (time, 0 min; IMD0). The regional intragastric meal distribution (IMD) was assessed using the methods of Parkman et al,^17^ with the proximal and distal halves of the stomach equally divided along the midpoint of the longitudinal axis of the stomach as previously described by Orthey et al and Piessevaux et al.^6,18^ For discrimination of impaired and normal fundic accommodation, 0.568 was used as the cut-off, as previously determined on receiver-operating-characteristic analysis.^6^

#### Gastric Alimetry

BSGM was performed using the Gastric Alimetry system, which includes a high-resolution stretchable electrode array (8×8 electrodes; 20 mm inter-electrode spacing; 196 cm^2^), a wearable Reader, an iPadOS App for concurrent validated symptom logging during the test.^9,12,19^ Array placement was preceded by shaving if necessary, and skin preparation (NuPrep; Weaver & Co, CO, USA). Recordings were performed simultaneously with GES encompassing 30 min fasting baseline, 10 min meal, and 4 h postprandial recording. Patients sat in a reclined relaxed position with limited movement, then transferred to the nuclear medicine table for imaging, with motion artifacts automatically corrected or rejected using validated algorithms.^20^ Symptoms including early satiation, nausea, bloating, upper gut pain, heartburn, stomach burn, and excessive fullness were measured during continuously testing at 15-minute intervals using 0-10 visual analog scales (0 indicating no symptoms; 10 indicating the worst imaginable extent of symptoms) and combined to form a ‘Total Symptom Burden Score’.^12^

BSGM spectral analysis included Principal Gastric Frequency (PGF; reference intervals: 2.65-3.35 cycles per minute), BMI-adjusted amplitude (reference intervals: 22-70 µV), and Gastric Alimetry Rhythm Index (GA-RI; reference intervals: ≥0.25) for BSGM.^21^ In addition, the ‘Meal Response Ratio’ (MRR) was used to characterise meal response timing; calculated as the ratio of the average amplitude in the first 2 hours postprandially to that of the last 2 hours.^11^ MRR was not calculated if postprandial recording duration was <4 h. A normal MRR was empirically defined as >1 based on previous studies,^21–23^ meaning that the dominant gastric motor response occurred within the first two hours after a meal. Phenotyping was based on an established consensus hierarchical approach:^24^

a. Neuromuscular phenotype: GA-RI <0.25 and/or BMI-adjusted amplitude <22
b. High frequency phenotype: PGF >3.35 cycles per minute
c. Delayed meal response phenotype: MRR <1 (considered normal spectrogram per current normative reference interval criteria)^21^
d. Normal phenotype: GA-RI, BMI-adjusted amplitude, and PGF within normative reference intervals

### Data analysis

All analyses were performed in Python v3.9.7 and R v.4.4.2 (R Foundation for Statistical Computing, Vienna, Austria). Continuous data were summarized as mean (standard deviation) or median (interquartile range) based on visual and statistical evaluation for normality, with appropriate tests for parametric or non-parametric data performed. Assumptions for correlation tests were assessed using the *performance* package with Spearman or Pearson coefficients reported accordingly.^25^ Categorical data were cross-tabulated, and differences tested using χ2 tests. To assess differential symptom impacts by results of multimodal physiological testing, a linear regression analysis was performed with interaction terms between Gastric Alimetry phenotypes and accommodation status based on IMD0. Results of linear regression are reported as β coefficient and 95% confidence intervals (CI).

## Results

67 patients underwent simultaneous gastric mapping and GES with sufficient data for meal distribution analysis (19 [28.4%] in Auckland and 48 [71.6%] in Western Sydney). Median age was 40 (IQR 28-56), 56 (84%) were female, and median BMI was 24 (23-30). The cohort is summarised in **Table 1**. On gastric emptying testing, median T1/2 was 54.5 (IQR 37.6-89.4), median percentage retained at 4 hours was 3% (IQR 0.7-11) with 18 (26.9%) meeting criteria for delayed emptying. The median IMD0 was 0.76 (IQR 0.69-0.86) with 5 (7.5%) classified as having impaired accommodation. Gastric Alimetry phenotyping showed 10 (14.9%) had a neuromuscular phenotype (low rhythm stability and/or amplitude), 5 (7.5%) had a high frequency, 18 (26.9%) had a delayed meal response ratio, and 34 (50.7%) had normal phenotype.

**Table 1:**
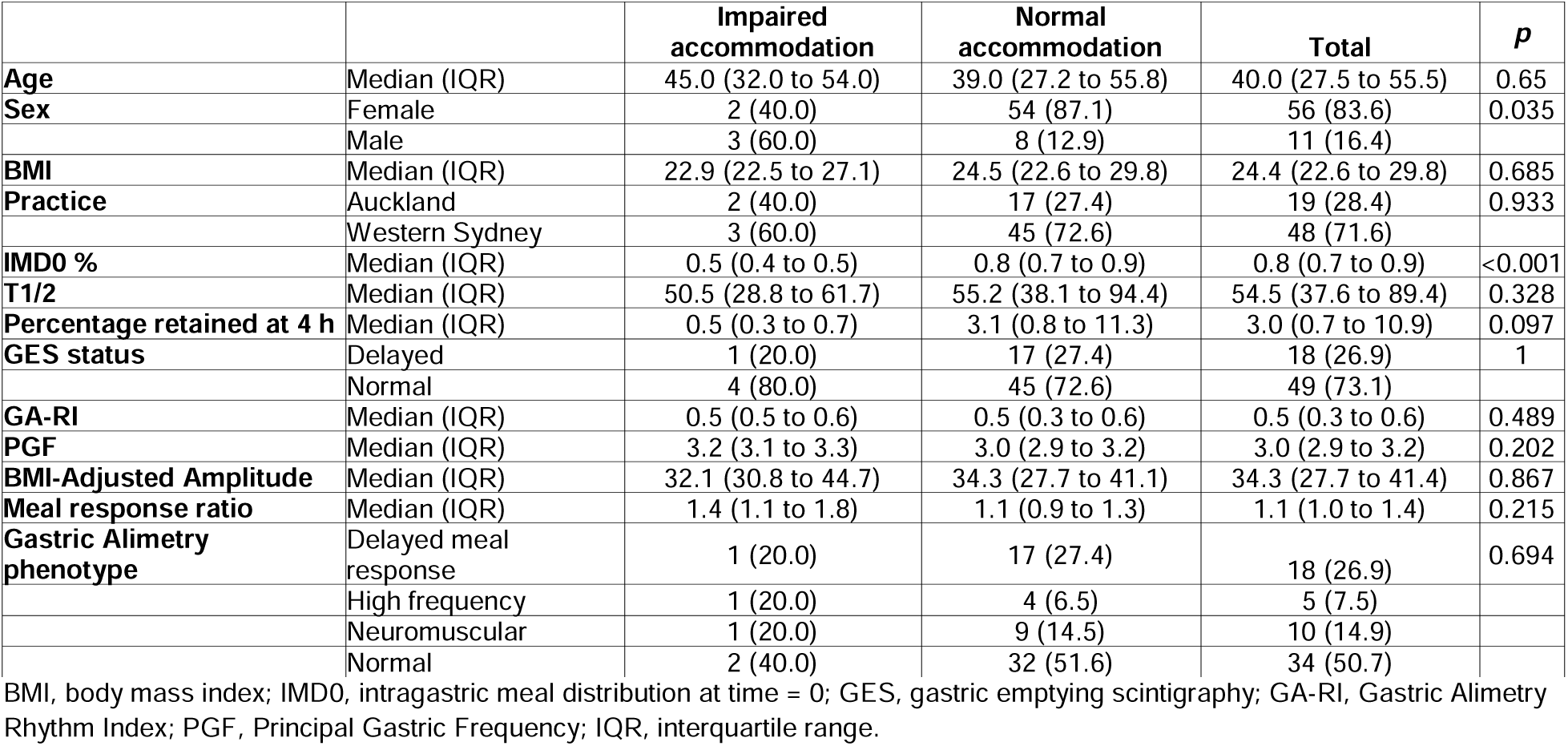
Demographic and physiological parameters stratified by accommodation status.

|  |  | **Impaired accommodation** | **Normal accommodation** | **Total** | **p** |
| --- | --- | --- | --- | --- | --- |
| **Age** | Median (IQR) | 45.0 (32.0 to 54.0) | 39.0 (27.2 to 55.8) | 40.0 (27.5 to 55.5) | 0.65 |
| **Sex** | Female | 2 (40.0) | 54 (87.1) | 56 (83.6) | 0.035 |
|  | Male | 3 (60.0) | 8 (12.9) | 11 (16.4) |  |
| **BMI** | Median (IQR) | 22.9 (22.5 to 27.1) | 24.5 (22.6 to 29.8) | 24.4 (22.6 to 29.8) | 0.685 |
| **Practice** | Auckland | 2 (40.0) | 17 (27.4) | 19 (28.4) | 0.933 |
|  | Western Sydney | 3 (60.0) | 45 (72.6) | 48 (71.6) |  |
| **IMD0 %** | Median (IQR) | 0.5 (0.4 to 0.5) | 0.8 (0.7 to 0.9) | 0.8 (0.7 to 0.9) | <0.001 |
| **T1/2** | Median (IQR) | 50.5 (28.8 to 61.7) | 55.2 (38.1 to 94.4) | 54.5 (37.6 to 89.4) | 0.328 |
| **Percentage retained at 4 h** | Median (IQR) | 0.5 (0.3 to 0.7) | 3.1 (0.8 to 11.3) | 3.0 (0.7 to 10.9) | 0.097 |
| **GES status** | Delayed | 1 (20.0) | 17 (27.4) | 18 (26.9) | 1 |
|  | Normal | 4 (80.0) | 45 (72.6) | 49 (73.1) |  |
| **GA-RI** | Median (IQR) | 0.5 (0.5 to 0.6) | 0.5 (0.3 to 0.6) | 0.5 (0.3 to 0.6) | 0.489 |
| **PGF** | Median (IQR) | 3.2 (3.1 to 3.3) | 3.0 (2.9 to 3.2) | 3.0 (2.9 to 3.2) | 0.202 |
| **BMI-Adjusted Amplitude** | Median (IQR) | 32.1 (30.8 to 44.7) | 34.3 (27.7 to 41.1) | 34.3 (27.7 to 41.4) | 0.867 |
| **Meal response ratio** | Median (IQR) | 1.4 (1.1 to 1.8) | 1.1 (0.9 to 1.3) | 1.1 (1.0 to 1.4) | 0.215 |
| **Gastric Alimetry phenotype** | Delayed meal response | 1 (20.0) | 17 (27.4) | 18 (26.9) | 0.694 |
|  | High frequency | 1 (20.0) | 4 (6.5) | 5 (7.5) |  |
|  | Neuromuscular | 1 (20.0) | 9 (14.5) | 10 (14.9) |  |
|  | Normal | 2 (40.0) | 32 (51.6) | 34 (50.7) |  |

BMI, body mass index; IMD0, intragastric meal distribution at time = 0; GES, gastric emptying scintigraphy; GA-RI, Gastric Alimetry Rhythm Index; PGF, Principal Gastric Frequency; IQR, interquartile range.

### Multimodal Physiological Assessment

Delayed gastric emptying (n=18) was associated with higher IMD0 (median 0.9 [IQR 0.8-0.9] vs 0.7 [IQR 0.6-0.8], p=0.004), indicating greater proximal retention after meal ingestion, and higher IMD0 also correlated with slower gastric emptying T1/2 (Pearson R=0.33, p=0.007; **Figure 1**). There was no significant difference in IMD0 between the different Gastric Alimetry phenotypes (p=0.4; **Figure S1**).

**Figure 1:**
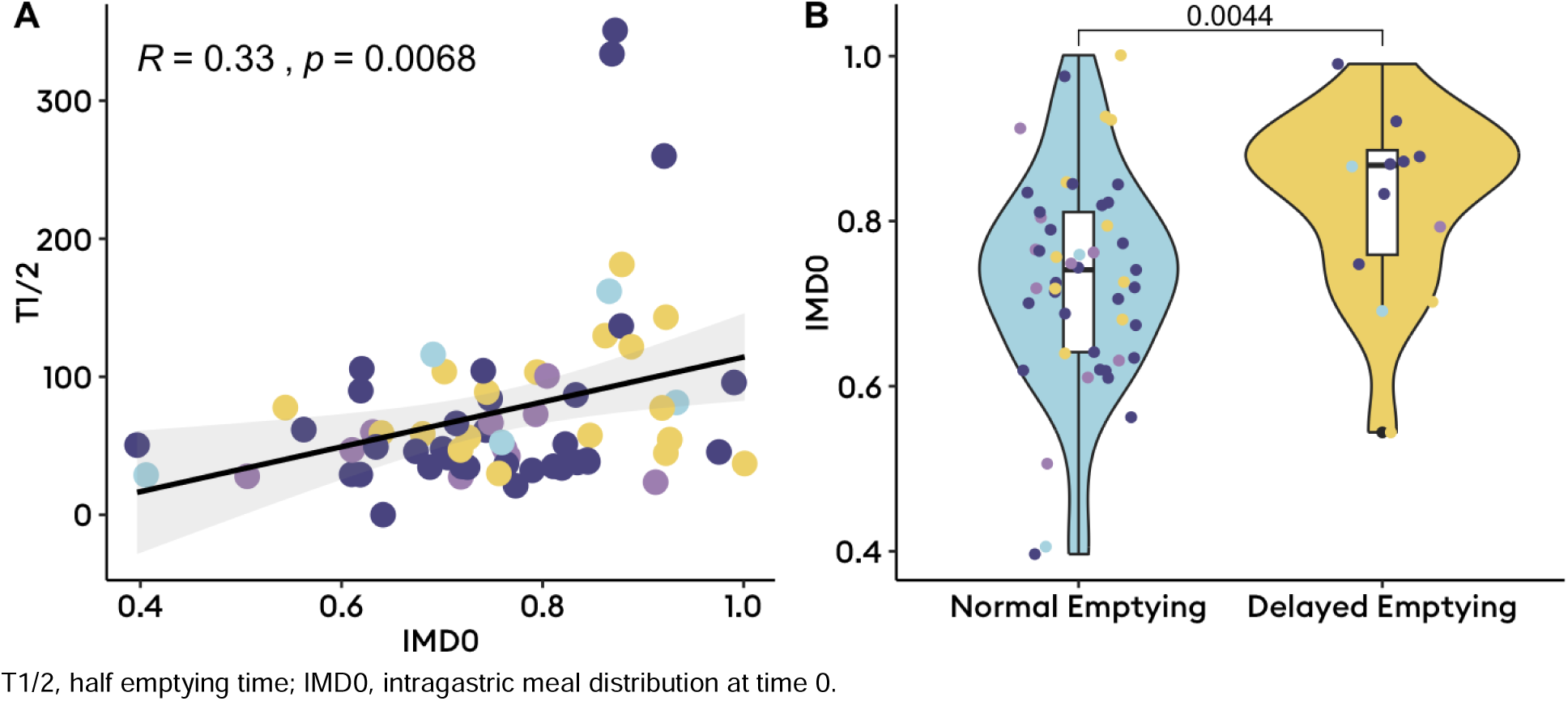
Relationship between gastric emptying scintigraphy transit delays and intragastric meal distribution. A: Correlation between T1/2 and intragastric meal distribution at time 0 (IMD0); B: Violin and box plots depicting the median, interquartile range, range and distribution of IMD0 by emptying status.

Among the 15 patients that had abnormal spectrograms (5 [7.5%] with high frequencies; 10 [14.9%] with low rhythm stability and/or amplitude), high IMD0 (proximal retention) was strongly correlated to delayed BSGM meal-response timing (Pearson R=-0.71, p=0.003; **Figure 2**) meaning proximal retention delayed the onset of gastric motor activity. This relationship was not seen among those with normal spectrograms (**Figure 2C-D**). Additionally, lower IMD, indicating antral distribution, was correlated with higher gastric frequencies (Spearman R=-0.27, p=0.03; **Figure 3**).

**Figure 2:**
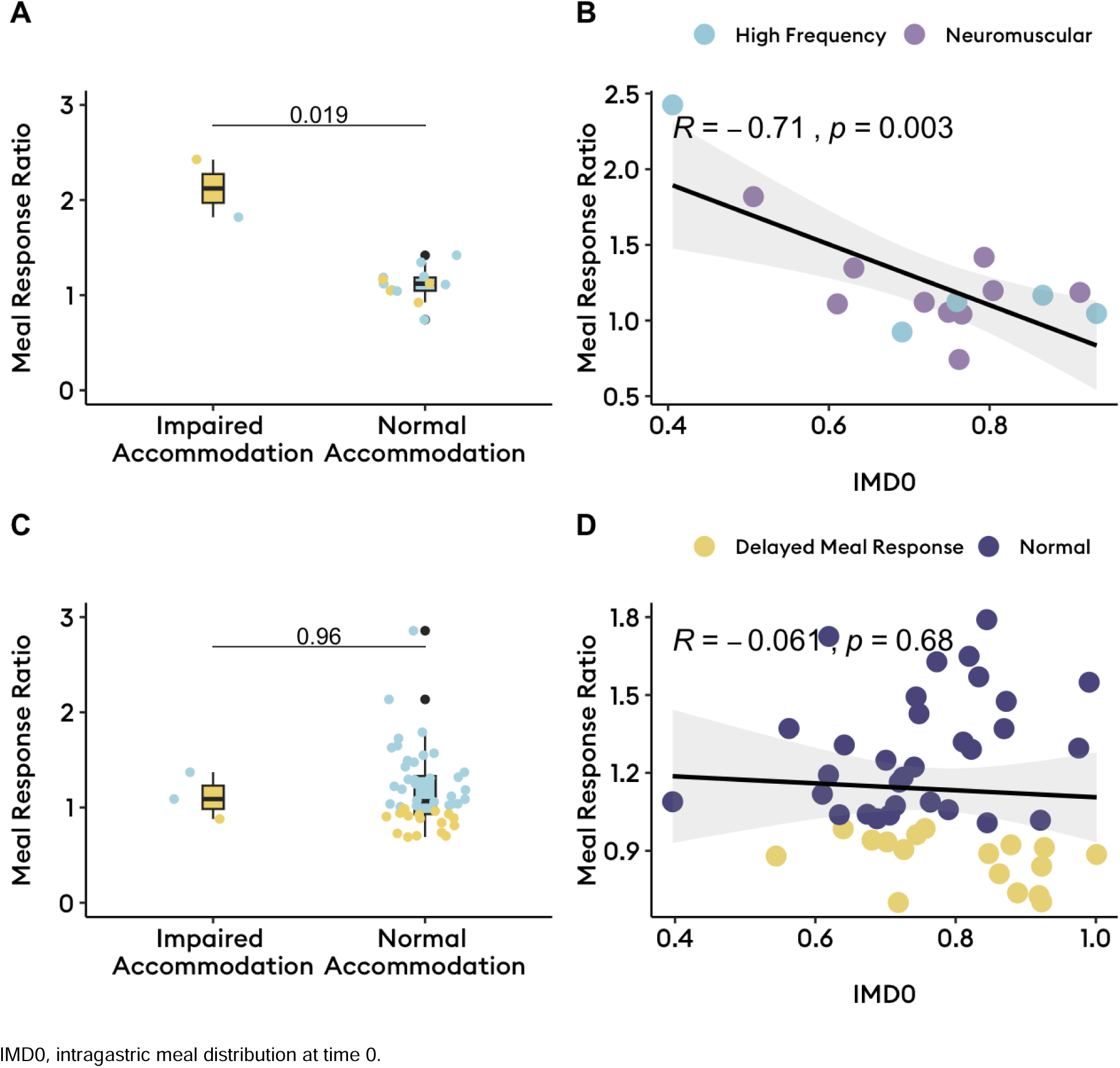
Impact of accommodation on meal response ratios and differential impacts by neuromuscular function. A-B: In those with abnormal spectrograms (n=18), impaired accommodation was associated with earlier meal responses (p<0.02); C-D: In those with normal spectrograms (n=52), meal distribution was not associated with meal response timing (p>0.65).

**Figure 3:**
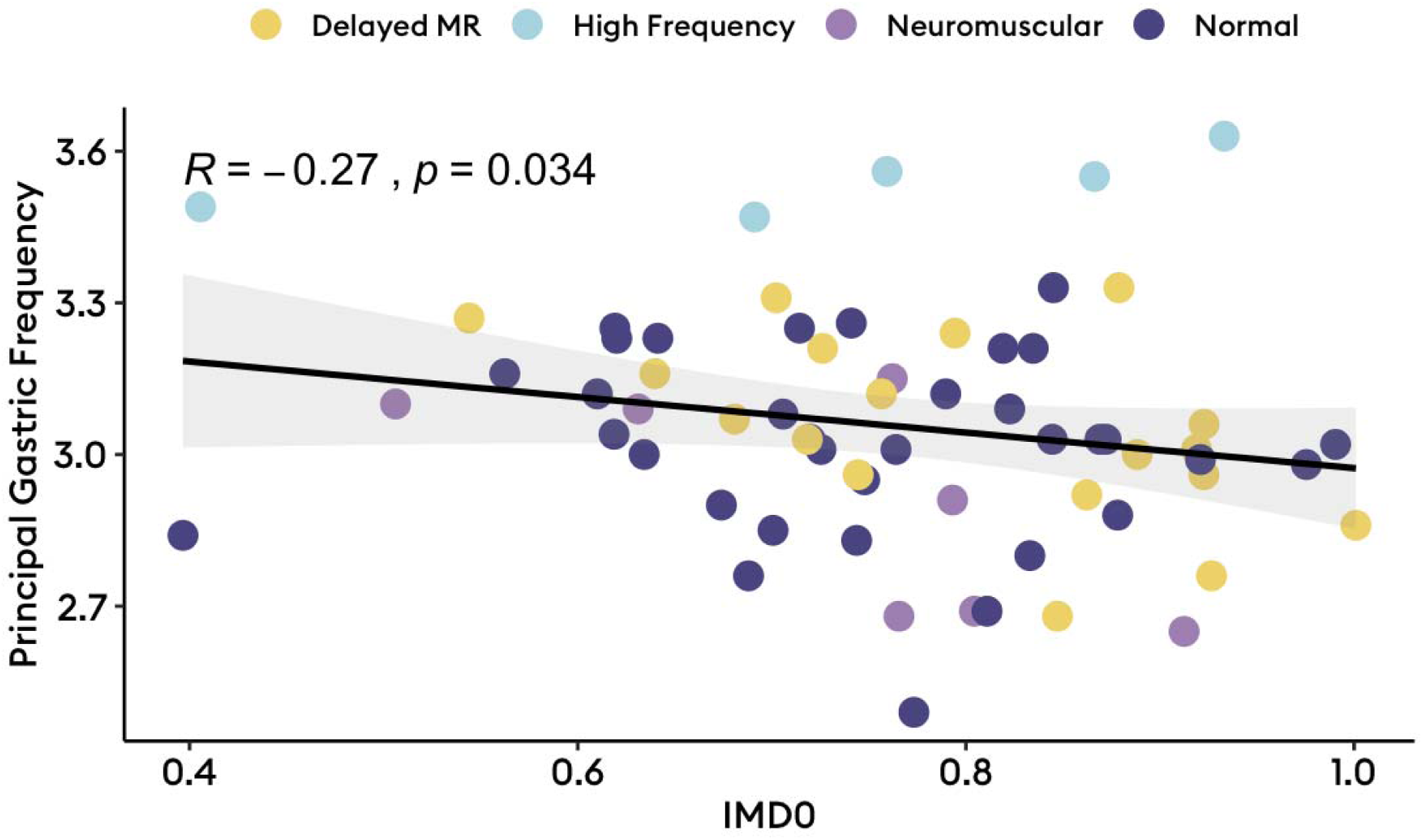
Correlation between frequency and intragastric meal distribution, with dots coloured by Gastric Alimetry phenotype

### Symptom Associations

Patients with a delayed meal response on BSGM had higher excessive fullness than those with a normal phenotype (mean difference 2.0, p=0.05). In addition, patients with a neuromuscular phenotype had lower heartburn symptoms than those with a normal phenotype (mean difference -2.1, p=0.04). Using a linear regression model, evaluating accommodation status by BSGM phenotype showed that the group with impaired accommodation on average had lower total symptom burden (β=-17, 95% CI -12 to -10, p<0.001), however when impaired accommodation was paired with a physiological abnormality on gastric mapping, total symptom burden was higher relative to those with normal IMD and BSGM (β>10, p<0.03; **Table 2**). Specifically, neuromuscular phenotype paired with impaired accommodation was associated with worse stomach burn (β=1.6, 95% CI 0.3-3.0, p=0.02) and excessive fullness (β=3.1, 95% CI 0.5-5.6, p=0.02); and high frequency phenotype paired with impaired accommodation was associated with worse bloating (β=4.5, 95% CI 2.3-6.8, p<0.001), excessive fullness (β=3.8, 95% CI 0.5-7.1, p=0.03), and stomach burn (β=4.4, 95% CI 2.6-6.1, p<0.001). Delayed meal responses paired with impaired accommodation was associated with worse upper gut pain (β=3.8, 95% CI 2.1-5.4, p<0.001), heartburn (β=2.8, 95% CI 1.4-4.2, p<0.001), stomach burn (β=1.8, 95% CI 0.5-3.2, p=0.009), and excessive fullness (β=2.7, 95% CI 1.0-4.4, p=0.002).

**Table 2:**
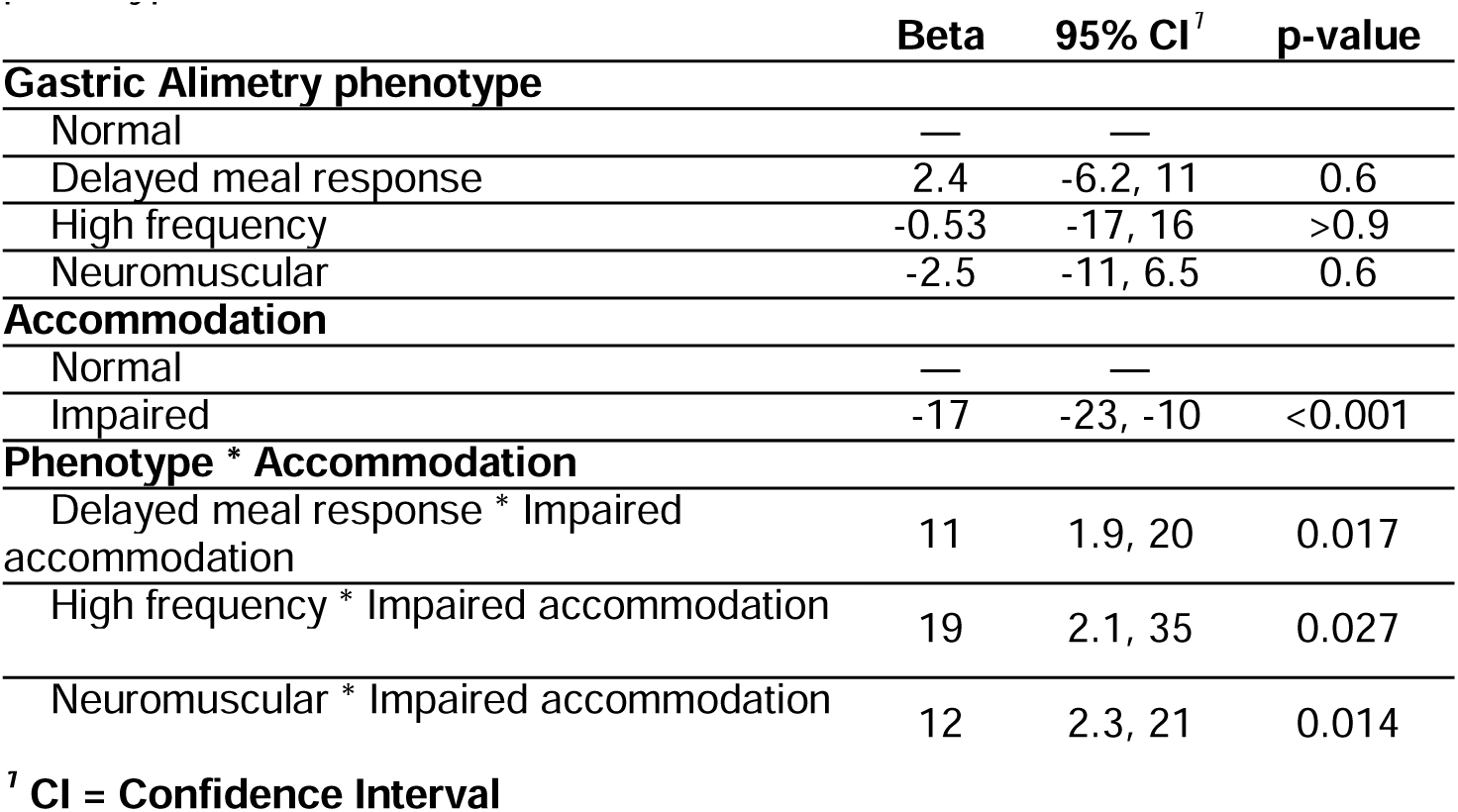
General linear model with interaction term between accommodation status and Gastric Alimetry.

## Discussion

This study applied multimodal physiological testing to patients with chronic gastroduodenal symptoms. We found that intragastric meal distribution influenced gastric emptying and postprandial motor responses on BSGM, in association with dyspeptic symptoms. Proximal gastric retention was associated with delayed emptying, and in the setting of abnormal gastric neuromuscular function, delayed meal responses. Impaired accommodation also most impacted those with abnormal spectrograms, indicating neuromuscular pathology, with antral predominant meal distribution inducing earlier postprandial antral motor responses that correlated with worse postprandial symptoms. Earlier postprandial antral motor responses also had an apparent chronotropic effect, with gastric frequency showing a mild negative correlation to IMD0. Notably, stomach motility was robust to aberrant measures of accommodation when neuromuscular function was intact (normal Gastric Alimetry spectrograms).

It has long been acknowledged that impaired fundic accommodation,^8^ and impaired postprandial antral motor activity are features of chronic gastroduodenal disorders.^26–28^ Troncon et al first demonstrated the relationship between impaired fundic accommodation and earlier redistribution of food to the antrum in patients with functional dyspepsia,^8^ suggesting it is the resultant premature luminal distension of the antrum that causes symptoms of bloating and fullness. The multimodal non-invasive physiological testing applied here, confirms these findings in individuals with underlying gastric neuromuscular dysfunction which confers reduced resilience to antral distension from impaired accommodation, causing dyspeptic symptoms, and simultaneously altering the gastric meal response.

Several factors influence slow wave chronotropy,^1^ importantly, including stretch. Mechanosensitivity in interstitial cells of Cajal,^29^ results in increased slow wave frequencies with distension, as supported in the present study by tendency toward higher gastric frequencies with more antral meal distributions (**Figure 3**). Excessive stretch, particularly in a neuromuscularly-impaired stomach may also predispose to decoupling of longitudinally propagating slow waves, impeding transit and inducing gastroduodenal symptoms.^30^ BSGM offers non-invasive biomarkers of gastric motor function, emphasising the clinical and physiological sequelae of fundic motor dysfunctions to the mechanosensitive ICC syncytium.^31,32^

This multicentre study used simultaneous scintigraphic and non-invasive gastric mapping in patients with chronic gastroduodenal symptoms. Strengths of this study included consistent IMD evaluation by a single author across the multiinstitutional dataset, simultaneous multimodal testing, and its reasonably large sample size, supporting the generalisability of these insights. However, this study has some limitations. Firstly, determination of impaired fundic accommodation is based on a consensus-based cut-offs of the IMD analysis which has not yet been externally validated.^6^ Further physiological validation using barostat or single photon emission computed tomography may offer added insights on impacts of gastric accommodation. Further, the therapeutic significance of paired myoelectrical and accommodation dysfunctions remains to be evaluated, with future therapeutic trials awaited.

In conclusion, multimodal physiological testing of gastric function shows that proximal gastric retention is associated with both delayed emptying and delayed postprandial motility responses on BSGM. Impaired accommodation had the most significant clinical impacts when myoelectrical abnormalities were present, with multiple motor dysfunctions on these tests being associated with worse dyspeptic symptoms. The addition of Gastric Alimetry BSGM biomarkers to standard functional assessment of the stomach aids in identifying gastric motility abnormalities.

## Data Availability

Patient-level data are not available. Group-level data are available on request.

## Acknowledgements

N/A

## Supplementary Appendix

**Figure S1:**
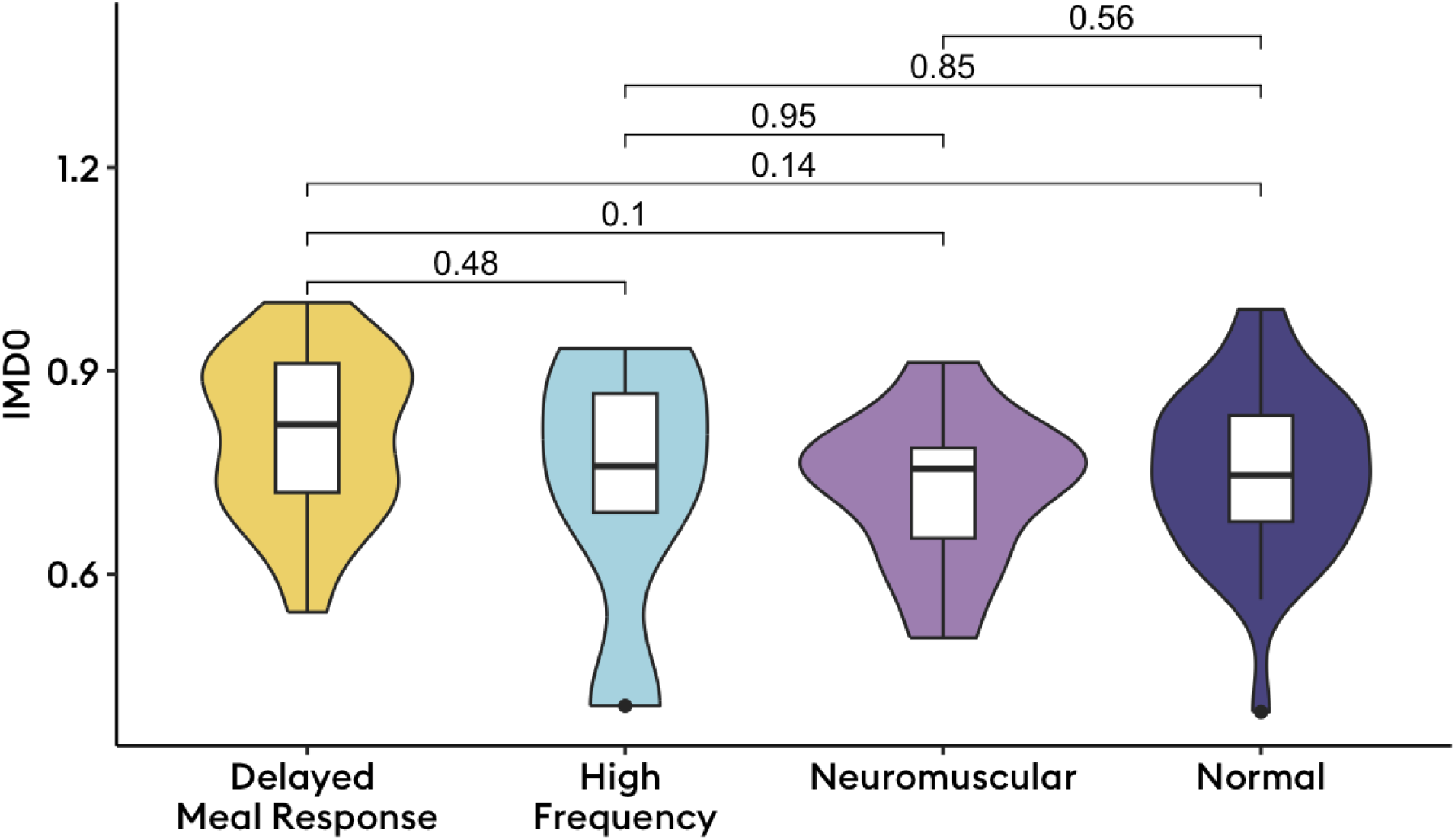
Intragastric meal distribution by Gastric Alimetry phenotype

## References

1. O’Grady, G., Gharibans, A. A., Du, P. & Huizinga, J. D. The gastric conduction system in health and disease: a translational review. American Journal of Physiology-Gastrointestinal and Liver Physiology 321, G527–G542 (2021).

2. Camilleri, M. Functional dyspepsia: mechanisms of symptom generation and appropriate management of patients. Gastroenterol. Clin. North Am. 36, 649–64, xi–x (2007).

3. Drossman, D. A. & Tack, J. Rome Foundation Clinical Diagnostic Criteria for Disorders of Gut-Brain Interaction. Gastroenterology 162, 675–679 (2022).

4. Pasricha, P. J. et al. Characteristics of patients with chronic unexplained nausea and vomiting and normal gastric emptying. Clin. Gastroenterol. Hepatol. 9, 567–76.e1–4 (2011).

5. Pasricha, P. J. et al. Functional Dyspepsia and Gastroparesis in Tertiary Care are Interchangeable Syndromes With Common Clinical and Pathologic Features. Gastroenterology 160, 2006–2017 (2021).

6. Orthey, P. et al. Intragastric Meal Distribution During Gastric Emptying Scintigraphy for Assessment of Fundic Accommodation: Correlation with Symptoms of Gastroparesis. J. Nucl. Med. 59, 691–697 (2018).

7. Febo-Rodriguez, L., Chumpitazi, B. P., Sher, A. C. & Shulman, R. J. Gastric accommodation: Physiology, diagnostic modalities, clinical relevance, and therapies. Neurogastroenterol. Motil. 33, e14213 (2021).

8. Troncon, L. E., Bennett, R. J., Ahluwalia, N. K. & Thompson, D. G. Abnormal intragastric distribution of food during gastric emptying in functional dyspepsia patients. Gut 35, 327–332 (1994).

9. Gharibans, A. A. et al. A novel scalable electrode array and system for non-invasively assessing gastric function using flexible electronics. Neurogastroenterol. Motil. e14418 (2022).

10. O’Grady, G. et al. Principles and clinical methods of body surface gastric mapping: Technical review. Neurogastroenterol. Motil. 35, e14556 (2023).

11. Varghese, C. et al. Distinct subgroups in gastroparesis defined by simultaneous body surface gastric mapping and gastric emptying breath testing. medRxiv (2024) doi:10.1101/2024.11.21.24317043.

12. Sebaratnam, G. et al. Standardized system and App for continuous patient symptom logging in gastroduodenal disorders: Design, implementation, and validation. Neurogastroenterol. Motil. 34, e14331 (2022).

13. Vandenbroucke, J. P., et al. Strengthening the Reporting of Observational Studies in Epidemiology (STROBE): explanation and elaboration. PLoS Med. 4, e297 (2007).

14. Abell, T. L. et al. Consensus Recommendations for Gastric Emptying Scintigraphy: A Joint Report of the American Neurogastroenterology and Motility Society and the Society of Nuclear Medicine. J. Nucl. Med. Technol. 36, 44–54 (2008).

15. Wang, W. J. et al. Gastric Alimetry® expands patient phenotyping in gastroduodenal disorders compared to gastric emptying scintigraphy. Am. J. Gastroenterol. (2023) doi:10.14309/ajg.0000000000002528.

16. Huang, I.-H. et al. Meal effects on gastric bioelectrical activity utilizing body surface gastric mapping in healthy subjects. Neurogastroenterol. Motil. e14823 (2024).

17. Parkman, H. P. et al. Relationships among intragastric meal distribution during gastric emptying scintigraphy, water consumption during water load satiety testing, and symptoms of gastroparesis. Am. J. Physiol. Gastrointest. Liver Physiol. 325, G407–G417 (2023).

18. Piessevaux, H., Tack, J., Walrand, S., Pauwels, S. & Geubel, A. Intragastric distribution of a standardized meal in health and functional dyspepsia: correlation with specific symptoms. Neurogastroenterol. Motil. 15, 447–455 (2003).

19. Gharibans, A. A. et al. Gastric dysfunction in patients with chronic nausea and vomiting syndromes defined by a novel non-invasive gastric mapping device. medRxiv (2022) doi:10.1101/2022.02.07.22270514.

20. Calder, S. et al. An automated artifact detection and rejection system for body surface gastric mapping. Neurogastroenterol. Motil. 34, e14421 (2022).

21. Varghese, C. et al. Normative Values for Body Surface Gastric Mapping Evaluations of Gastric Motility Using Gastric Alimetry: Spectral Analysis. Am J Gastroenterol 118, 1047–1057 (2023).

22. Gharibans, A. A. et al. A novel scalable electrode array and system for non-invasively assessing gastric function using flexible electronics. Neurogastroenterology & Motility 35, e14418 (2023).

23. Gharibans, A. A. et al. Gastric dysfunction in patients with chronic nausea and vomiting syndromes defined by a noninvasive gastric mapping device. Sci. Transl. Med. 14, eabq3544 (2022).

24. Varghese, C. et al. A standardized classification scheme for gastroduodenal disorder evaluation using the gastric alimetry system: Prospective cohort study. Gastro Hep Adv. (2024) doi:10.1016/j.gastha.2024.09.002.

25. Assessment of Regression Models Performance [R package performance version 0.13.0]. (2025).

26. Rees, W. D., Miller, L. J. & Malagelada, J. R. Dyspepsia, antral motor dysfunction, and gastric stasis of solids. Gastroenterology 78, 360–365 (1980).

27. Camilleri, M., Malagelada, J. R., Kao, P. C. & Zinsmeister, A. R. Gastric and autonomic responses to stress in functional dyspepsia. Dig. Dis. Sci. 31, 1169–1177 (1986).

28. Malagelada, J. R. & Stanghellini, V. Manometric evaluation of functional upper gut symptoms. Gastroenterology 88, 1223–1231 (1985).

29. Won, K.-J., Sanders, K. M. & Ward, S. M. Interstitial cells of Cajal mediate mechanosensitive responses in the stomach. Proc. Natl. Acad. Sci. U. S. A. 102, 14913–14918 (2005).

30. Chan, C.-H. A. et al. Localized gastric distension disrupts slow-wave entrainment leading to temporary ectopic propagation: a high-resolution electrical mapping study. American Journal of Physiology-Gastrointestinal and Liver Physiology 321, G656– G667 (2021).

31. Mercado-Perez, A. & Beyder, A. Gut feelings: mechanosensing in the gastrointestinal tract. Nat. Rev. Gastroenterol. Hepatol. 19, 283–296 (2022).

32. Servin-Vences, M. R. et al. PIEZO2 in somatosensory neurons controls gastrointestinal transit. Cell 186, 3386–3399.e15 (2023).

