## Supplementary material for "Relationship between intragastric meal distribution, gastric emptying and gastric neuromuscular dysfunction in chronic gastroduodenal disorders": Table 2

**Table 2:** General linear model with interaction term between accommodation status and Gastric Alimetry phenotype

|  | **Beta** | **95% CI***^1^* | **p-value** |
| --- | --- | --- | --- |
| **Gastric Alimetry phenotype** |  |  |  |
| Normal | — | — |  |
| Delayed meal response | 2.4 | -6.2, 11 | 0.6 |
| High frequency | -0.53 | -17, 16 | >0.9 |
| Neuromuscular | -2.5 | -11, 6.5 | 0.6 |
| **Accommodation** |  |  |  |
| Normal | — | — |  |
| Impaired | -17 | -23, -10 | <0.001 |
| **Phenotype * Accommodation** |  |  |  |
| Delayed meal response * Impaired accommodation | 11 | 1.9, 20 | 0.017 |
| High frequency * Impaired accommodation | 19 | 2.1, 35 | 0.027 |
| Neuromuscular * Impaired accommodation | 12 | 2.3, 21 | 0.014 |
| ***^1^* CI = Confidence Interval** | | | |
